# Effectiveness of a psychosocial training intervention in reducing psychological distress among caregivers of intellectually disabled children in Malawi – a randomized wait-list trial

**DOI:** 10.1101/2022.04.11.22273704

**Authors:** C. Masulani-Mwale, F. Kauye, M Gladstone

## Abstract

**Background:** Rates of disability are high in resource-poor settings with eighty-five percent of disabled children living in these settings. Long-term caregiving for disabled children is associated with fatigue, financial difficulties, parenting distress, and other psychological issues. Studies have shown a link between parenting children with intellectual disabilities and psychological distress as well as overall Health-Related Quality of Life (QoL). However, with interventions, these negative impacts may not be as severe as thought before. This study aimed at developing and testing the impact of a contextualized psychological intervention, Titukulane, in reducing psychological distress among caregivers with intellectually disabled children in Malawi.

**Methods:** We conducted a randomized waitlist trial of a psychosocial training intervention (Titukulane) provided to caregivers of children with intellectual disabilities. Caregivers of children with intellectual disabilities aged 0 to 18 years were recruited, screened, and then enrolled in the trial through two disability organizations operating in Mzuzu (St John of God) and Lilongwe (Children of Blessings). They were then randomized in blocks to the Titukulane intervention or waitlist and provided with the intervention or standard care for 3 months respectively. Assessment of socioeconomic status, age, gender, and maternal psychological distress (through the Self Reported Questionnaire (SRQ) were conducted at baseline and follow-up.

**Results:** We found that psychological distress on SRQ was significantly lower in caregivers of children with intellectual disability in the Titukulane intervention in comparison to the waitlist control group even when the confounding variables of age, gender, and social-economical status were taken into account (Cohen d = 0.08; CI = 0.33-0.754; p =0.0005).

**Conclusions:** Pyschosocial interventions such as the Titukulane intervention provided over a few months can improve caregiver mental health and quality of life – an important factor for supporting families of children with intellectual disability.

## Introduction

The diagnosis of intellectual disability is becoming more prevalent as we are becoming more aware and more able to diagnose it in LAMIC settings (1, 2). Intellectual disability is characterized by an impairment of skills manifested during the developmental period contributing to difficulties in the overall level of intelligence, i.e., cognitive, language, motor, and social abilities (3). Despite the increased awareness of this as a diagnosis in LAMIC settings, studies have shown that many caregivers suffer from stress, family issues, and the inability to cope with children with these difficulties (4-6). Presently, there are very few robust psychological interventions to alleviate these issues in most developing countries, particularly in Africa (7, 8).

Several theoretical perspectives appear to be influential in describing the relationship between the psychological impact of caring for an intellectually disabled child and the effect of the psychosocial interventions in addressing that impact. Some models include the ABC model, the Family Care Giving model, and Risk-resilient model. The Family Care Giving Model is a model which provides support in understanding two concepts: parenting and parents’ psychological health. This model which uses the theory of stress and coping theory by McCubbin identifies three major stages in stress and coping: antecedents, mediators, and outcomes (9). The antecedents considered in some studies are child characteristics (functional dependency, the severity of a disability and caregiver or family characteristics (parent education, income, and parent or family functioning). The mediators are some of the interventions discussed above. Categories of adaptation outcomes hypothesized can include caregiver’s improved coping skills and reduced psychological distress, culminating in improved quality of life; and child’s living status and participation in age-appropriate community activities. This model is considered influential as it highlights the detailed mechanisms of action for parenting and its effects on parental psychological health. The limitation of this model, however, is that McCubbin has advanced a negative stance on the impact of parenting and the psychological health of the parents of these intellectually disabled children. The other theoretical perspective is the Risk-resilient model proposed by Wallander which accounts for the adaptation of parents of intellectually disabled children with disabilities (10). This model contains risk factors (parameters of child’s disability, parents’ functional care strain, and parents’ psychosocial stress) and resistance or resilient factors of the parents that alleviate the stressors (coping, support, and social-ecological factors). The limitation of this model is that it presumes that parenting disabled children has negative consequences on parental psychological health and hence considers it a risk always when it can also have positive consequences in the long run. The fact that one study has shown that long-term parenting may lead to a reduction in parental stress (8), leaves an unanswered question on whether the psychosocial interventions are the only factor responsible for the improved psychological health outcomes in the parents or if there are other factors?

The development and trialing of effective treatments for use by non-specspecialists are among the top research priorities for improving the mental health of caregivers of children with disabilities in the developing world (11). Although there are many RCTs done to test Psychosocial Training Interventions in the West, there have been few such RCTs, and those that have, come from Asia (12). In Malawi, there are no published trials of interventions aiming to support caregivers for children with disabilities, and in particular, none which aim to support the mental health of the caregivers. This paper describes a wait-list RCT that was undertaken to trial the Titukulane intervention for reducing psychological distress among caregivers with intellectually disabled children in Malawi.

### Study objective

The objective of this study was to determine the effectiveness of Titukulane, compared to the usual health education, in reducing psychological distress among caregivers with intellectually disabled children.

#### Null hypothesis

The Psychosocial Training Intervention (Titukulane) is not effective in reducing psychological distress among caregivers with intellectually disabled children than the usual health talk.

## Methods

### Study design

We conducted a wait-listed randomized trial as a rigorous technique to evaluate Titukulane’s short-term impact on maternal mental health.

### Study population, screening, and enrolment

Participants in this study were caregivers of children with intellectual disabilities aged 0 to 18 years in clinics in Mzuzu and Lilongwe. They were recruited after their children met set screening criteria for intellectual disability through the use of the ten-question questionnaire (TQQ) and then secondarily, a screen by a psychiatric clinical officer who confirmed the DSM-5 criteria which included:

1) that the child had a deficit in intellectual functioning—”reasoning, problem-solving, planning, abstract thinking, judgment, academic learning, and learning from experience”—confirmed by clinical evaluation and individualized standard IQ testing using Wechsler Intelligence Scale for Children (WISC) (13); and 2) a deficit in adaptive functioning that significantly hamper conforming to developmental and sociocultural standards for the individual’s independence and ability to meet their social responsibility (14).

### Randomization

The study recruited caregivers of children with intellectual disabilities who were able to give informed consent. Caregivers whose children had intellectual disabilities were consecutively allocated to the study teams and allocated to the two arms of the study through block randomization with blocks of five in one group and five in the other for every ten consecutively referred caregivers and either the second phase (15). The allocation sequence was computer generated by the Principal investigator. To ensure concealment of the allocation sequence, sequentially numbered, opaque, sealed envelopes were used.

### Blinding

Although randomization minimizes differences between treatment groups at the outset of a trial, it does nothing to prevent differential treatment of the groups later in the trial or the differential assessment of outcomes which may result in biased estimates of treatment effects. This study, therefore, used double-blinding where both investigator/assessors and participants are unaware of allocation. The interventions (Titukulane versus usual education talks) were similar in terms of delivery because they were all administered as group education interventions.

### Sample Size

To calculate a sample size for this study, three factors were considered as follows: 1) The level of statistical significance, normally 5% (16); 2) the power, that is the probability to reach statistical significance) for a given effect size, was set at 90% (17); and an effect size of 0.4 based on analysis by Singer et.al., for a similar intervention (18). The calculated sample size, therefore, was 162 (comprising 81 participants needed for each arm of the study). We further factored in a 10% loss to follow-up by adding 8 subjects to each group for a total sample size of 89 in each arm.

### Data collection tools and data collection

At both baseline as well as at a three-month follow-up, we verbally administered the study questionnaire to all consenting participants in quiet rooms that provided privacy for about 15 to 20 minutes. We collected demographic details of the participants which are associated with psychosocial distress. These included center/site (Mzuzu vs Lilongwe); Age which in a subsequent was redefined into a dichotomous variable of vulnerability (Vulnerable = too young/too old to parent [these would struggle to care for the child due to young age or old age] versus less vulnerable= middle-aged parents [these were are productive and can provide care]), (Young versus old): Social economic status (Lower versus Upper SES) through wealth rankings; Gender (Female versus Male); Occupation (Unemployed versus Employed); Education (Up to primary versus Secondary & above); Marital status (Never married versus Married); Knowledge of one’s child’s disability (No Vs Yes): and Availability of support [availability of counseling or accompaniment received from professionals, and others who are supportive of caregivers’ mental health concerns, and based on Protégé–mentor agreement psychosocial support by Noe’s 14 item-Psychosocial Support mentoring scale of 1988] (19). We also asked about the caregivers’ knowledge of their child’s disability (20).

We also collected data using the Self-Reporting Questionnaire (SRQ) as the measure of psychological distress. The SRQ is a quick measure of mental health problems designed by the WHO to be used across cultures (21). It has been validated, in Malawi, against Structured Clinical Interview for DSM-IV (SCID) for major depressive episodes in mothers of infants attending measles vaccination in Thyolo District Hospital (22). In this Malawian study, SRQ had high internal consistency (Cronbach’s alpha 0.85], and the area under the ROC curve for detection of current major depressive disorder was 0.856 (22). Our study, therefore, adopted the 7/8 cut-off-point because of good epidemiological indices for major/minor depression. The SRQ is scored out of 20. Participants who scored 8 and above were considered distressed while those scoring 7 and below were considered non-distressed (20). While SRQ was the primary outcome, four sub-factors of psychological distress using the factor analysis adopted from the weights from the WHO SRQ User’s Guide manual were computed (23). This was done using principal component factor analysis with varimax rotation for the psychological distress score (23). These included: decreased energy, somatization, depressive mood, and depressive thoughts.

### The Titukulane intervention

After baseline screening, caregivers were exposed to the Titukulane (intervention group) or usual health education (control group) depending on randomization. A detailed description of the Titukulane intervention is described in our previous study (24) however, in summary, the intervention is a “contextualized psychosocial intervention for parents with intellectually disabled children” which consists of ten-40 minutes sessions conducted twice every week. Participants were requested to attend all training sessions and transport cost to the venues was reimbursed after the sessions. All research assistants were trained in the administration of the questionnaires with emphasis on the measurement of all research outcomes above and the ethical considerations of the study.

### Data management and analysis plan

Data analysis was performed with STATA version 13.0 software (25). The effect analysis for this study was based on a modified intention to treat analysis or explanatory analysis. All participants who had missing data on their independent variable (SRQ) or dropped from the study were excluded from the final analysis.

Assessment of baseline characteristics was done using proportions and is presented in Table 6.1. Baseline characteristics were compared using the Chi-squared test (χ2) and T-tests. For this study, both univariate and multivariable analyses were conducted using Odds Ratios and linear regression models respectively. Two-sided P-values were used and confidence intervals were calculated at the 95% level. Statistical significance was set at P ≤ 0.05. Moderator analysis was done to check if the nature and strength of the relationship between the main outcome variable (psychological distress) and one explanatory variable (like social-economic status) changed as a result of a function of an explanatory.

The four sub-factors of psychological distress from the SRQ were analyzed using factor analysis adopted from the WHO SRQ User’s Guide manual using principal component factor analysis with varimax rotation for the psychological distress score (SRQ). These were analyzed as secondary outcomes within the trial. For this analysis, we utilized WHO weights for the four sub-factors as they are based on a large group of 1182 participants. The SRQ subfactors included; Factor I-Decreased energy or lethargy; Factor II-Somatic symptoms; Factor III-Depressive mood; and Factor IV-Depressive thoughts.

### Ethical Considerations

Great care was taken to ensure that all ethical issues as detailed in the Helsinki Declaration were followed. All subjects are treated with respect and provided the right to refuse participation in the study as well as to have the interviews conducted in privacy. Permission to carry out the study was sought from the two institutional heads for St John of God and Children of Blessings Trust; ethical clearance was sought from the COMREC (# P.06/14/1591); while written informed consent was sought from all participants before participation in the study. The study was also registered with ClinicalTrials.gov with ID: NCT02827396

## Results

### Enrollment and baseline

Out of 265 caregivers who were assessed for eligibility in this study, 7 declined to consent procedures, and 67 were not eligible (because their child was aged above 18 years; they had parented the child for less than a year, or the caregivers were aged less than 18 years. Four declined because they intended to move out of the study area, while one moved out of the area before randomization. A total of 186 caregivers were randomized to the intervention or control group. (Refer Figure 1). Of the 95 participants allocated to intervention, 94 had complete data sets at baseline screening however of the 91 control participants, 13 did not attend the trial sessions. These were not significantly different from the control sample. (Refer figure 1 Below). There was good adherence to the intervention as all participants attended all sessions in both arms.

**Figure 1.**
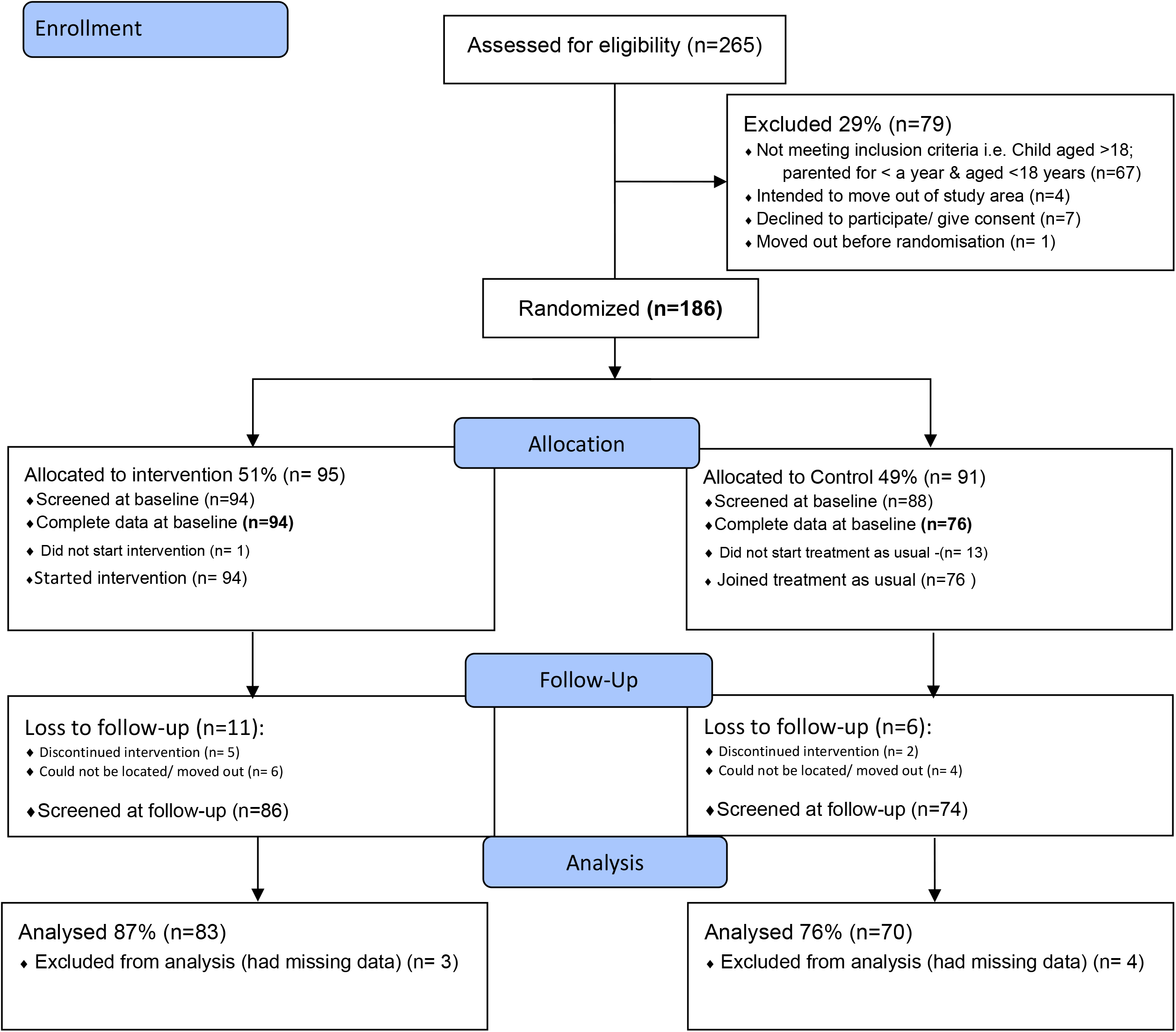
Study flow diagram for the RCT for Titukulane intervention for parents of children with intellectual disabilities

At follow-up, 11 of the intervention group participants were lost to follow-up; five dropped out and discontinued the intervention, and six could not be located leaving 86 participants who were screened at the three-month follow-up. Six control participants were lost to follow-up; two dropped out and four could not be located. This left 74 participants who were screened at the three-month follow-up.

Three participants in the intervention arm and four participants in the control arm were excluded from the analysis due to missing data (Figure 1).

### Baseline social-demographic characteristics of participants

The mean age of participants was 34.4 years (SD: 9.5). The majority of participants were female 90.1 % (n=156) with only 39 (21%) of the participants not married. The majority of participants; 103 (55.7%) had completed secondary education; with a surprisingly high number; 64 (34.6%) having completed their tertiary education. A large percentage, (62.4%) of the participants, were unemployed (n=116), while those who were in employment (n=70). 38% (n=70) of the participants had high socioeconomic status, while 17.4% and 44.6% had middle or low socioeconomic status respectively. Many participants (68.3%) indicated that they had no psychological support. This included; no availability of counseling or accompaniment received from professionals, religious and community leaders who are supportive of participants’’ psychological and mental health concerns, and respond appropriately as needed (19); while 59 (31.7%) had the required support. Knowledge of one’s child’s disability, very few caregivers did not know the nature of their child’s disability. Just less than half; 47.3% (n=88) of caregivers were from Lilongwe while the rest were from Mzuzu.

### Pre- and Post-intervention Prevalence of Psychological distress

At baseline, the prevalence of psychological distress (SRQ of more than or equal to 7) overall in both groups combined was 41% (20) with the intervention group having 39.4% of caregivers with high psychological distress and the control group, having 43%.

At follow-up, the prevalence of psychological distress among the intervention group had dropped to 7.2% while the rate in the control group increased to 47% (Figure 2). This was statistically significant (χ^2^= 31.8; α= 0.0005). Mean SRQ scores for psychological distress in the intervention and control groups were 4.95 (CI: 4.37 - 5.53) and 7.18 [CI: 6.56 - 7.8) respectively (t= 5.1; p value= 0.0005) with a reduction in scores on SRQ in the intervention group in comparison to the control group at the time of follow-up.

**Figure 2:**
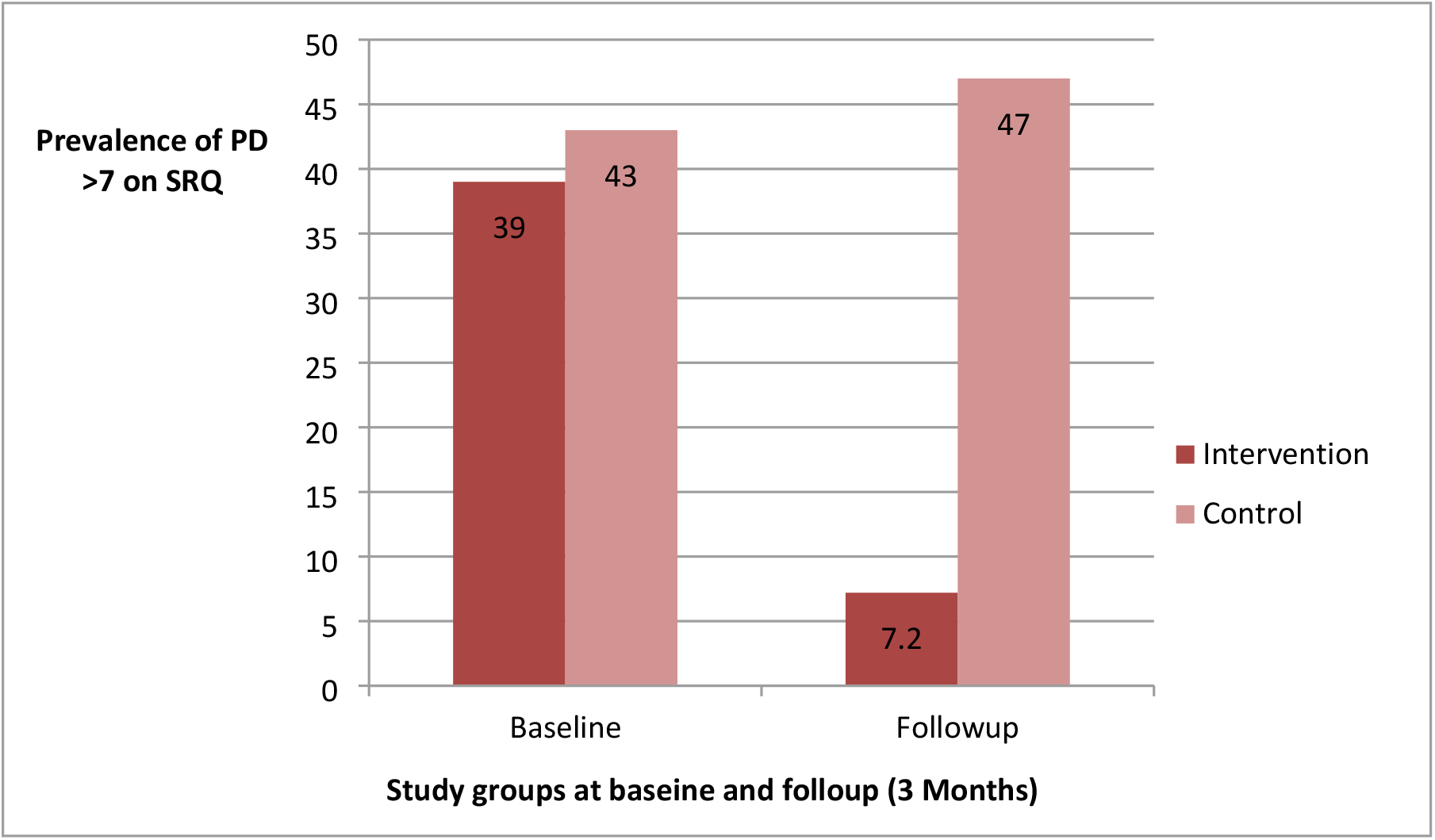
Figure demonstrating Pre- and Post-intervention prevalence of psychological distress (PD) among participants in the Titukulane intervention study

### The effect size of the Titukulane on psychological distress of caregivers

In addition to our statistical analysis above, we computed an effect size for the Titukulane intervention which showed that the intervention had a medium practical significance effect (Cohen *d* =0.08; CI = 0.33-0.754; p =0.0005).

### Predictors of psychological distress and impact of the Titukulane

Bivariate analysis showed that there were significant differences between the intervention group and the control group in the risk of psychological distress with psychological distress in the intervention group much lower than in the control group (Crude Odds Ratio=0.08; CI= 0.03 to 0.2; p =0.0005).

We found the two following factors to be associated with a reduced risk of psychological distress (Table 2) 1) Higher socio-economic status, and 2) Already having psychological support (OR=0.3; CI= 0.1 - 0.7, and 0.35; CI = 0.1-0.9) respectively). The risk of psychological distress among Lilongwe participants did not fall as much as those who were living in Mzuzu (OR=0.30; CI= 0.16 to 0.7; p-value =0.001).

**Table 2:** Showing psychological distress and associated factors assessed in the two groups of participants enrolled in the intervention at follow-up.

| Variable |  | Psychological distress |  | Crude Odds Ratios |  | Adjusted Odds Ratios |  |
| --- | --- | --- | --- | --- | --- | --- | --- |
|  | Category | Positive | Negative | Crude OR (CI) | P-value | Adjusted OR (CI) | P-value |
| Location | Mzuzu(*Reference*) | 27 (35.1%) | 50 (64.9%) |  |  |  |  |
|  | Lilongwe | 12 (15.8%) | 64 (84.2%) | 0.3 (0.16-0.7) | 0.006* | 0.5 (0.2-1.2) | 0.12 |
| Age | Young and old aged (*Reference*) | 32(25%) | 96 (75%) |  |  |  |  |
|  | Middle aged | 7 (28%) | 18 (72%) | 0.85 (0.32-2.2) | 0.99 | 0.99 (0.95-1.05) | 0.95 |
| Socio-economic status | Low SES(*Reference*) | 26 (38.8%) | 41 (50%) |  |  |  |  |
|  | Upper SES | 13 (15.8%) | 73 (84.9%) | 0.2 (0.13-0.6) | 0.001* | 0.3 (0.1-0.7) | 0.01* |
| Sex/Gender | Female(*Reference*) | 3 (23.1) | 11 (76.9%) |  |  |  |  |
|  | Male | 35 (25.4%) | 103 (74.6%) | 1.07 (0.3-3.5) | 0.91 | 1.3 (0.35-5.2) | 0.65 |
| Occupation | Unemployed(*Reference*) | 28 (29.5%) | 67 (70.5%) |  |  |  |  |
|  | Employed | 11 (19%) | 47 (81%) | 0.56 (0.2-1.23) | 0.148 | 0.9 (0.37-2.3) | 0.87 |
| Education | Up to primary(*Reference*) | 31 (22.5%) | 107 (77.5%) |  |  |  |  |
|  | Secondary & above | 8 (53.3%) | 7 (46.7%) | 0.25 (0.08-0.75) | 0.09 | 0.35 (0.1-1.1) | 0.73 |
| Marital status | Never married(*Reference*) | 27 (22.3%) | 94 (77.7%) |  |  |  |  |
|  | Married | 12 (37.5%) | 20 (62.5%) | 0.47 (0.2-1.10) | 0.08 | 0.58 (0.23-1.4) | 0.25 |
| Knowledge of one's child's disability | No(*Reference*) | 8 (22.9%) | 27 (77.1%) |  |  |  |  |
|  | Yes | 31 (26.3%) | 87 (73.7%) | 1.2 (0.49-2.92) | 0.68 | 1.1 (0.4-2.9) | 0.84 |
| Psychological Support | No support(*Reference*) | 33 (55.9%) | 26 (44.1%) |  |  |  |  |
|  | Has support | 32 (30.5%) | 73 (69.5%) | 0.38(0.15-0.96) | 0.036* | 0.35 (0.1-0.9) | 0.03* |
| Recent Emotional event | No(*Reference*) | 24 (20.5%) | 93 (79.5%) |  |  |  |  |
|  | Yes | 21 (58.3%) | 15 (41.7%) | 2.7 (1.2-6.1) | 0.01* | 2.5 (1.04-6) | 0.04* |
| Group |  |  |  |  |  |  |  |
|  | Control(*Reference*) | 33 (84.6%) | 37 (32.5%) |  |  |  |  |
|  | Intervention | 6 (15.4%) | 77(67.5%) | 0.08 (0.03-0.2) | 0.0005* | 0.75 (0.02-0.3) | 0.0005* |

\* Adjusted for location, Social-economic status, source of psychological support, and occurrence of a recent emotional event.

We found significant differences between participants who had recently experienced an emotional event (e.g. death of a loved one) and those who did not. Participants who reported to have experienced an emotional event were 2.7 times more likely to have psychological distress than those who had not experienced such an event (OR=2.7; CI= 1.2 to 6.1; p-value = 0.01). Other variables (age, sex/gender’ occupation, education, marital status, and knowledge of one’s child’s disability) were not significantly associated with psychological distress.

After multivariable logistic regression analysis, we continue to see a reduced risk of psychological distress in the intervention group, however, the effect of the intervention is less pronounced (Adjusted Odds Ratio of 0.75; (CI= 0.02 to 0.21; p =0.0005).

Multivariable logistic regression analysis, after controlling for all variables significant in the univariate analyses (location, social-economic status, source of psychological support, and occurrence of recent emotional event), suggests that participants who reported to have experienced an emotional event were 2.5 times more likely to have psychological distress than those who had not experienced such an event (OR=2.5; CI= 1.04 to 6); p-value = 0.04). Belonging to higher socio-economic groups and having psychological support availability were protective from psychological distress (OR=0.3, CI: 0.1-0.7 and OR=0.35, CI: 0.1-0.9 respectively). The other variables (location, age, socio-economic status, sex/gender occupation, education, marital status, and knowledge of one’s child’s disability) were not significantly associated with psychological distress.

### Moderator analysis

We aimed to conduct moderator analysis with the two strongest co-variates (socio-economic status and psychological support) where regression models were compared before and after adding these two computed moderator variables into the model. There was no significant moderation with the percentage of psychological distress explained by the two variables remaining at 10% before and after including these moderator variables into the model (Adjusted R^2^ =0.10 before adding a moderator variable and Adjusted R^2^ =0.103 after including a moderator into the model). The two variables were not statistically significant in explaining psychological distress in both models.

### Factor analysis of SRQ scores for psychological distress

On analysis of the sub-factors, participants in both groups scored highly on lethargy, depressive mood, and depressive thoughts, but very few on somatization. The control group scored higher than the intervention group for every factor but particularly for “depressive mood”. (Refer figure 3).

**Figure 3:**
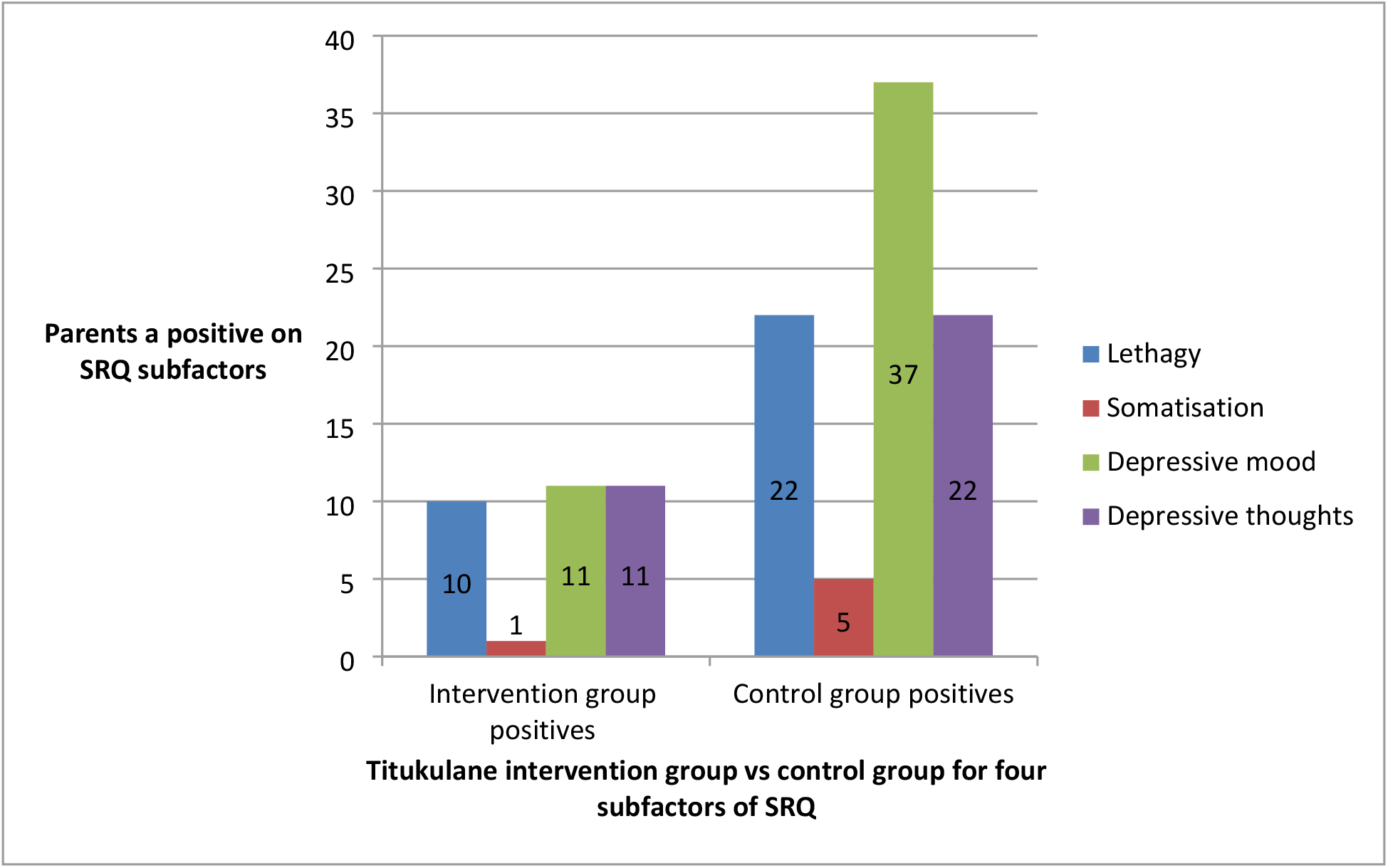
Demonstrating percentage of caregivers scoring positive in each of the WHO SRQ subfactors in the Titukulane intervention group vs control

### Impact of Titukulane on Lethargy (Decreased energy); somatization; Depressive mood and

At follow-up, there were significant differences between the intervention group and the control group in the risk of decreased energy. Decreased energy was reduced in the intervention group relative the control group (Crude Odds Ratio=0.2; CI= 0.1-0.5; p-value =0.001).

When somatization was used as a dependent variable in bivariate analysis, no statistically significant difference was found in somatization, between participants in the intervention group and the control group (OR=0.15; CI= 0.12-1.3; p value=0.58).

On depressive mood, there was a high risk for depressive mood among participants in the control group than those in the intervention group (Crude Odds Ratio =0.3; CI =0.6-0.29; p-value =0.0005). Further, the risk of depressive thoughts was reduced among those with upper socioeconomic status and those with secondary and tertiary education relative to those participants from low socioeconomic status and those with only up to primary education respectively (OR =0.3, and 0.2 respectively).

## Discussion and conclusion

Our results support the efficacy of carers’ intervention training on mental health for caregivers of children with intellectual disabilities in Central Africa. We have demonstrated through our wait-list block-randomized controlled trial, a definite statistically significant fall in the percentage of carers of children with intellectual disability scoring highly (> 7) on the SRQ when provided with the Titukulane intervention. The adjusted Odds Ratio above and Cohen’s effect size value (*d* = 0.70) suggest a large to very large substantive practical effect size for the developed intervention. We have demonstrated that this works and is effective even when associated co-variates are considered. The caregivers in the intervention group benefited from the intervention compared to those in the control group. We have demonstrated that this intervention supports caregivers in many ways but particularly with depressive symptoms. This fall in rates of poor mental health, in particular depressive symptoms, even when being provided with a short intervention, is very encouraging. Our study is comparable to development work in Pakistan where a Tablet-based android application was developed for training, monitoring, and supervision of the champions, based on the World Health Organization’s guidelines. The champions delivered the intervention to other families in the network (26). The project was sustainable and demonstrated significant improvements in the lives of children and their families, in the first 6 months of its operation. Our current study only measured the impact of Titukulane at 3 months follow-up. The findings are also comparable to evaluations of group-based parent training programs for caregivers of children with developmental disabilities, which show that psychological intervention leads to improvements in mental health outcomes (27-29).

This study’s impact has been higher than that shown in other similar studies. This high impact may be explained by the fact that the Titukulane was multidimensional; and that it was the first of its kind to be administered among such caregivers. The fidelity to the intervention can be attributed to cultural acceptability and relevance of the intervention, particularly on how it was created in a ground-up culturally relevant way using the most recent evidence (24). It is clear that many caregivers of children with intellectual disabilities have marked features of depression and mental health issues in Malawi – it is likely this is one reason that it worked well.

Further to this, in our sites, no such robust intervention is offered so it may have been an extremely helpful opportunity for caregivers, hence the huge impact. Group-based interventions have also shown effects of increasing perceived social support for participants (30). This is because the group meetings served as a source of peer learning and encouragement to caregivers that struggle with their parenting role and hence this may lead to the huge effect size seen in this intervention.

This huge impact is similar to other studies which have shown that psychosocial interventions bring positive mental health outcomes among caregivers of intellectually disabled children (31-34). The large effect size can be explained by the fact that the program had good attendance and a low attrition rate of less than 15%. The impact may also be because transport was paid for. It was also a quite intensive program over a short duration and that’s why we observed such huge effects immediately but which may not be so much later after a large period of follow-up say in 9 to 12 months. This is similar to trends in other studies or for similar interventions (35). In addition to these findings, the high impact could also be explained by the fact that caregivers liked the intervention. The qualitative data in the intervention quality assessment pilot study showed great satisfaction with the Titukulane.

While this study had a high impact, some studies have found minimal to no effect of group-based intervention compared to individual interventions (36, 37). The minimal effectiveness in these negative findings has been associated with poor acceptability and limited intervention’s emphasis on mental health outcomes In our study, however, many mothers were suffering from poor mental health, and therefore, it may be the case that any intervention might have been beneficial. Our research demonstrates this to be the case, but more may need to be done to understand which factors within the program and what fidelity is needed to enable this to be maintained if scaled up. Some studies have also demonstrated that while there was preliminary evidence of efficacy in reducing negative child behaviors related to group intervention, no post-intervention change in depression among carers of disabled children was found. This has been attributed to the fact that the group training does not emphasize practical interventions for stress and depression (38). Concern was also raised that Behavioral Parent Training (BPT) may exacerbate parental stress by placing additional demands and expectations on the caregivers (39). On the other hand, for caregivers with more resilience, research has shown that they may already be perceiving benefits and positive contributions associated with caring for their disabled children, and hence the interventions may not have any added post-intervention impact (40).

### Factors associated with psychological distress

It was found that living in Mzuzu; belonging to an upper socioeconomic level, better parent education, and already having psychological support were associated with reduced psychological distress. Much of these factors could be related to the economic circumstances of the population. In Mzuzu, the poverty headcount is less than that of Lilongwe (41). Secondly, individuals with good socio-economic status and better parent education are more likely to seek and receive better support both materially and psychologically. This may also be due to an increased capacity to travel and visit centers, to understand child disability issues and seek the required support, and hence have less unmanaged psychological burden resulting in low psychological distress among participants belonging to these classes.

There were more female caregivers than males who took part in our study. In Malawi, women are still the main caregivers for children. Furthermore, single motherhood is more common in mothers of children with disabilities often as a result of marital breakdown caused by the birth of the disabled child in the family (42, 43). This gender bias in the burden of care of all children with disabilities is evident (43), even when considering other main caregivers from the extended family. In future studies, it may be useful to consider how we might understand more about how this may influence the mental health of mothers.

In this present study the majority of participants sampled were unemployed and from a low socioeconomic status; which is often interweaved within this is the fact that those mothers are therefore more likely to have infants and children with disabilities. Secondly, many poor mothers are less likely to access a health service due to the costs associated with it.

It was clear from the present study that those who had recently experienced an emotional event like the death of a loved one were associated with much psychological distress. It would be assumed that this can be explained by the stress associated with the loss which results in psychological distress (44-46).

What was more concerning was that knowledge of one’s child’s disability was associated with much psychological distress. This could have resulted in caregivers focusing much thought and attention on their child’s disability and its impact rather than focusing on some positive aspects of this role as has been reported in some studies (47, 48). These thoughts and preoccupations could be building up, and end up exacerbating distress symptoms among the caregivers (38).

### Study limitations and strengths

While the intervention was highly effective, the conclusions drawn from the data need to be treated with caution due to some of the methodological challenges encountered during the study. The first limitation was the low sample size of 89 participants per arm. This may have affected the power and the results of the study. Like in Most Randomised Wait-List trials, the substantial drop-out of participants at follow-up may have also affected the overall conclusions of the study (49). This attrition can influence the balance of confounders between the groups influence the statistical power of the study and its ability to detect an effect of the intervention if it exists. Therefore, the positive outcomes may have been favorable for these caregivers who participated in the intervention and completed the outcome measures for the study. The second limitation may be that the participants could have been underreporting symptoms which is a common issue in self-reports. It should be noted /that SRQ scores may have been so good at follow-up because of issues surrounding blinding and self-reports which were used in collecting data from the participants. This study only used double-blinding and underreporting by participants may not be ruled out completely. The use of parents’ self-report of their psychological distress may not be the most reliable measure of psychological distress due to over or under-reporting by the participants (50, 51). Related to this could be that there was a positive bias whereby caregivers were over satisfied with the intervention leading to underreporting of distressing symptoms after the training (52). Thirdly, other possible confounding variables (like the severity of child disability and number of disabled children per parent e.t.c) were not measured. Measuring these would have helped to assess if such moderating factors had an impact on the effect of the intervention.

Fourthly, the high impact of these findings cannot be guaranteed for a long period of follow-up because it was only done three months post-intervention. As detailed below, there may be a need to conduct a study to ascertain if protection can be maintained at 9 months and 12 months post-intervention, for example.

Lastly, the generalization of these findings may be limited to urban settings and surrounding environs because the study did not sample participants from the very rural population.

Despite these shortcomings, the strengths of this study suggest that the findings can be considered with confidence. Firstly this was a randomized study and as such it could give a more robust impact of the intervention. Well-validated and reliable instruments (TQQ and SRQ) were used in the screening and assessment protocol of the children and caregivers respectively (53). Secondly, the intervention was delivered by trained health workers. While the use of trained health care workers was good, the limitations of this concern the difficulty to replicate in the community at a larger scale, in the future, because we don’t have many trained personnel at the community level. The use of non-specialized personnel is being promoted in community settings (54).

And lastly, the intervention had multidimensional components namely: modules focusing on psychological support, social support, financial support, and the use of group experience sharing among others. This could have contributed to the success of the intervention.

### Implications for future research

While this study ascertained the efficacy of the Titukulane, future studies need to deepen such findings to investigate the mechanisms of change resulting from the intervention by using rigorous conceptual models (55). This would help to understand the mediators and moderators of change involved for two groups of participants. This understanding of the mechanisms of change would allow for greater prudence in intervention administration, easier roll-out to practical community settings, and ultimately optimization of therapeutic change. Secondly, future studies may need to measure other possible confounding variables like the severity of child disability and the number of disabled children per carer, to ascertain their effect on the reduction of psychological distress.

In our current study, we had high uptake, adherence, and fidelity with the intervention with quite controlled circumstances and well-trained people doing the intervention – this may have been a reason for the excellent results we had. Families were provided with transport to attend. This may be a factor that enabled good adherence and therefore good outcomes and would be important to explore in the future as to whether this could be effective in the same way if scaled up, possibly with fewer resources within community settings in Malawi. Another future question may be around whether it could be scaled up in the same way with more local staff around the country and what the fidelity and adherence at this level might be. In addition, there is a need for a longer period of follow-up studies, to check if this effect size and post-Titukulane protection can be maintained at 9 months and 12 months follow-up. Regarding clinical practice, the intervention has been demonstrated to be efficacious in reducing psychological distress among these caregivers, hence showing that it has to find a place in primary mental health care and disability services in the country and sub-region. There is also a need to build capacity among community health workers so that this intervention can be rolled into our community-based rehabilitation clinics to promote this work in the future.

## Data Availability

The data underlying the results presented in the study are available from the author upon request

## Declarations

### Consent for publication

Not applicable.

### Availability of data and materials

The datasets used and/or analyzed during the current study are available from the corresponding author on reasonable request

### Competing interests

The authors declare they have no competing interests.

### Funding

This study was supported by the Consortium for Advanced Research Training in Africa (CARTA). CARTA is jointly led by the African Population and Health Research Center and the University of the Wits and funded by the Wellcome Trust (UK) (Grant No: 087547/Z/08/Z), the Department for International Development (DfID) under the Development Partnerships in Higher Education (Delphi), the Carnegie Corporation of New York (Grant No: B 8606), the Ford Foundation (Grant No: 1100-0399), Google.Org (Grant No: 191994), Sida (Grant No: 54100029) and MacArthur Foundation Grant No: 10-95915-000-INP. These funders did not play any part in the design of the study and collection, analysis, and interpretation of data and in writing the manuscript.

## Acknowledgments

We are grateful for the following for their input in this paper: Glory Mwafuirwa and Napasyanga Nyondo who helped in data collection; the parents and children with disabilities; Mr. Nyirenda, the psychiatric C. M. O. who helped with the recruitment and screening the children; the clinic workers in Lilongwe and Mzuzu; Dr. Jupter Simbeye of University od Malawi who supported with data analysis.

## Author Contributions

▪ Concept and design of paper: CM
▪ Methodology: CM, MG, DM, and FK
▪ Discussion: CMs and MG
▪ Writing original draft: CM & PK
▪ Critical revisions of the paper: MG and DM
▪ Study supervisors: MG, DM and FK

## Notes

### Competing Interest Statement

The authors have declared that no competing interests exist.

### Clinical Trial

ClinicalTrials.gov with ID: NCT02827396

### Author Declarations

College of Medicine Research and Ethics committee as established in 2006 to support local researchers in designing and conducting clinical research to introduce ICH-Good Clinical Practice (GCP) quality research standards to provide comprehensive training in GCP, Research Methodology, Grant Proposal Writing, Statistical Analysis and Data Management and to manage research grants with the overall goal of developing a research agenda and climate that will allow sustainable and successful competition for international grants and local research that impact local health policy. My approval number # P.06/14/1591

